# “Epidemiological profile of patients with stage 5 chronic kidney disease on dialysis with Covid 19 infection in a Public Hospital-Perú”

**DOI:** 10.1101/2021.09.30.21264132

**Authors:** Yanissa Venegas-Justiniano, César Loza-Munarriz, Abdías Hurtado-Aréstegui

## Abstract

**Introduction:** Chronic kidney disease (CKD) in Covid 19 is relevant, however, there are few descriptions and fewer in Peru. Our goal was to describe the epidemiological profile and the factors related to mortality and survival of patients with stage 5 (CKD) on chronic dialysis hospitalized for Covid-19 in a public hospital.

**Methods:** Retrospective case series. Patients with stage 5 CKD, older than 18 years, hospitalized for Covid-19 infection were included. The primary data source was medical records. The clinical and epidemiological profile of the study sample and the factors related to mortality and survival are described.

**Results:** 105 medical records of patients with CKD 5 were evaluated. 57 (54,29%) were male, with a mean age of 58,59 ± 14,3 years. 84 (80%) patients survived and 21 (20%) died. The main cause of admission to hospitalization was respiratory failure in (80) 76,2%. The hospital stay was 11,76 ± 7,8 days. In the bivariate analysis: the increase in leukocytes, D dimer, ferritin, CRP, LDH and the decrease in lymphocytes, pH, bicarbonate and PaO_2_/FiO_2_ were related to mortality. In the multivariate analysis, only CRP> 10 mg/dl [HR: 10.72 (95% CI 1,4-81,58)] and a PaO_2_/FiO_2_ ≤ 150 mmHg [HR: 44,40 (95% CI 5,86-336,06)] they were factors related to poor survival.

**Conclusions:** CRP levels> 10mg/dl and PaO_2_/FiO_2_ ≤ 150 mmHg are the main factors related to mortality and poor survival in patients with stage 5 CKD hospitalized for Covid-19.

## INTRODUCTION

The World Health Organization (WHO) declared a pandemic to the infection for coronavirus 19 (COVID-19) in March 2020, initially this was identified in a group of patients with respiratory compromise in December 2019 in Wuhan. The infection spread quickly in Asia, Europe and the rest of the continents, in Peru, the first case was identified in March 6 of 2020 and to date numerous cases and a high mortality rate have been reported, putting it in the fifth most affected country in Latin America**^1-7^**.

According to reports, the infection is more frequent in the elderly population, men, comorbidities like diabetes mellitus and other immunosuppression conditions. The clinic presentation is known by sustained fever, lymphopenia, hyperferritinemia, pulmonary involvement in more than 50% of cases and other findings in relation to increased interleukins **^8-12^**.

Chronic kidney disease (CKD) characterized by the progressive loss of renal function, it has a progressive increase in frequency and the need of replacement therapy in any of its modalities is increasingly demanding **^2^**. The population with CKD both in pre-dialysis and renal replacement therapy, as well as in renal transplant recipients have multiple comorbidities associated with higher mortality during COVID-19 infection. It is known that the immune response of the chronic renal patient is diminished with less possibility of presenting cytokine storm, which explains why some series show a low percentage of severe manifestations. However, it should be considered that the chronic state of these patients, associated comorbidities, nutritional level and hospital admissions make them more prone to severe respiratory infections **^12-15^**.

Up to now, existing studies show different findings, some of them include that hemodialysis patients with COVID-19 experiment a minor illness with mild pneumonia, associated with a reduced immune response and decreased cytokine storm **^15,16^**. On the contrary, other series report a mortality of 28% in patients on hemodialysis, with hospital admission and severe clinical manifestations **^17^**. The most important finding is the fact that factors such as advanced age, diabetes, cardiovascular disease, pulmonary disease, and a less efficient immune system with the need for dialysis treatment in overcrowded environments, together with years of dialysis, lead to an increase in the prevalence and mortality rate **^18-21^**.

In Latin America and Peru, little has been described about the burden of Covid 19 infection in hospitalized patients with stage 5 chronic kidney disease on dialysis; characteristics, behavior, evolution and complications, these data are important to optimize care strategies and clinical outcomes.

The main objective of the study is to describe the epidemiological and clinical characteristics and factors related to mortality and survival of patients with stage 5 in chronic kidney disease on dialysis with Covid 19 infection in a public hospital in Lima-Peru.

## METHODS

The present study is a retrospective case series, conducted at the National Hospital Arzobispo Loayza (HNAL). Lima-Peru, between April and December 2020. The study population was selected by non-probabilistic sampling; hospitalized patients with stage 5 CKD on renal replacement therapy (RRT) and Covid-19 infection were evaluated.

Inclusion criteria were hospitalized patients older than 18 years, patients with stage 5 CKD in a chronic hemodialysis or chronic peritoneal dialysis program, renal transplant patients with functioning kidney; clinical diagnosis of Covid-19 infection by their treating physician and/or diagnosis of Covid-19 in the discharge summary. Exclusion criteria were patients with a diagnosis of Acute Kidney Injury (AKI), episode of AKI superadded to CKD and incomplete data in the clinical history records, database, or laboratory record. All were new cases of Covid 19.

During the study period, 107 patients with a diagnosis of stage 5 CKD and COVID 19 infection were registered; 105 met the inclusion criteria. The primary source of data was the medical records of each patient, and the data were recorded retrospectively, assigning sequential numbering, evaluated only by the investigator.

The independent variables considered for the study were: Age, sex, BMI, hypertensive and non-hypertensive etiology of CKD, symptomatologic profile on admission, smoking history, cause of hospitalization, comorbidities (Charlson Score), inflammatory parameters (CRP, ferritin, D-dimer, DHL, lymphocyte and leukocyte counts), blood gas values (pH, bicarbonate, pO_2_ and PaO_2_/FiO_2_), urea, creatinine, type of vascular access, hospital stay, renal replacement therapy (RRT) modality: chronic hemodialysis (HDC) or chronic peritoneal dialysis (CPD) and associated complications to COVID-19. Dependent variables were high vital status and survival time.

### Ethical Aspects

The Institutional Ethics and Research Committee of the Cayetano Heredia Peruvian University approved the study under the number 039-01-21 and since it is a retrospective study, it is exempt from informed consent.

### Statistical analysis

Descriptive statistics: Descriptive statistics were used to describe numerical variables with means ± SD for variables with normal distribution and with medians and interquartile range (IQR) for variables without normal distribution. Categorical variables were described in proportions (%). We report the crude mortality rate and frequency tables that include clinical and demographic characteristics, laboratory alterations and variables related to hospitalization, treatments, and complications of patients with stage 5 CKD on dialysis and Covid 19 infection. A survival analysis is realized presenting the curve and the general survival table of the study sample.

Inferential Statistics: Bivariate analysis was performed to compare clinical and demographic characteristics and laboratory data between survivors and deceased. Variables with a p ≤ 0.2 were selected to be evaluated in the multivariable model. To compare categorical variables or proportions, the “Chi2 exact” test was used. To compare two means of independent samples with normal distribution, the “ttest” (Student’s t-test) was used and to compare two means of independent samples without normal distribution, the “Wilcoxon rank sum” test was used. To compare more than two means for data with normal distribution, ANOVA and/or One-way was used.

The tables and the overall survival curve of patients with stage 5 CKD on dialysis and Covid-19 infection in the study period were obtained. Survival by groups was also described. The survival curves were compared with the Log Rank test. To evaluate the variables that were independently related to the overall survival of CKD stage 5 patients and Covid 19 infection, a multivariate analysis was performed with Cox regression using the Hazard Ratio (HR). The data were analyzed with State vs. 17 software. For the analysis, p ≤0.05 was considered statistically significant.

## RESULTS

In the study period were evaluated 105 patients with a diagnosis of stage 5 CKD and Covid-19 infection, the average age was 58.59 ± 14.3 years, 57 (54.29%) patients were male, respiratory failure 80 (76.2%) was the main cause of hospitalization; the most frequent etiology of CKD was arterial hypertension in 57 (54.28%) patients and diabetic nephropathy in 26 (24.76%); the hospital stay was reported as 11.76±7.8 days **(Table 1)**.

**Table 1:** General characteristic of the CKD5 dialysis and Covid 19 patients admitted to hospitalization.(n=105)

| Characteristic | N° | % |
| --- | --- | --- |
| **Male** | 57 | 54.29 |
| **Age** | 58.59±14.3 |  |
| **CKD etiology** |  |  |
| Nephroangiosclerosis | 57 | 54.28 |
| Diabetic nephropathy | 26 | 24.77 |
| Unknown | 16 | 15.25 |
| Obstructive uropathy | 3 | 2.85 |
| Others | 3 | 2.85 |
| **Cause of hospitalization** |  |  |
| Respiratory Insufficiency | 80 | 76.2 |
| Acid-base disorder/electrolytes | 11 | 10.47 |
| Severe anemia | 5 | 4.76 |
| Sensory disorder | 4 | 3.81 |
| Other | 5 | 4.76 |
| **Comorbidities** |  |  |
| Arterial hypertension | 102 | 97.14 |
| Diabetes mellitus | 25 | 23.8 |
| Obesity | 8 | 7.61 |
| Obstructive uropathy | 6 | 5.71 |
| Hypothyroidism | 4 | 3.81 |
| Others | 6 | 5.71 |
| **Income Symptom** |  |  |
| Dyspnoea | 70 | 66.67 |
| General discomfort | 13 | 12.38 |
| Fever | 12 | 11.42 |
| Other | 14 | 11.42 |
| **BMI** |  |  |
| Underweight | 1 | 0.95 |
| Normal | 64 | 60.96 |
| Overweight | 32 | 30.98 |
| Obese | 8 | 7.61 |
| **Death** | 21 | 20 |
| **Hospital stay duration(days)** | 11.76±7.8 |  |
| **Charlson Index** |  |  |
| 2 | 26 | 25.49 |
| 3 | 9 | 8.82 |
| 4 | 37 | 36.27 |
| 5 | 22 | 21.57 |
| 6 | 8 | 7.84 |

**BMI: body mass index; CKD5: chronic kidney disease stage 5**

The admission modality was as new in 40 (38.1%) and readmission in 65 (61.9%); pulmonary congestion 46 (43.81%), hyperkalemia 40 (38.1%), metabolic acidosis 12 (11.43%) and uremic encephalopathy 5 (4.76%) were the most frequent indications for dialysis; mostly received hemodialysis in 104 (99.04%) and only 1 (0.96%) peritoneal dialysis; the initial vascular access was a temporary central venous catheter 51 (48.57%), tunelled catheter 34 (32.38%) and 20 (19.05%) arteriovenous fistula; when evaluating the oxygen requirement 46 (43.81%) needed a oxigen mask and 39 (37.14%) a binasal cannula, an average BMI of 24.3±3.87 was reported.

In the bivariate analysis, the increase in leukocytes, D-dimer, ferritin, CRP, LDH, as well as the decrease in pH, bicarbonate, lymphocytes and PaO_2_/FiO_2_, were related to higher mortality **(Table 2 and 3)**.

**Table 2:** General characteristic of the CKD5 dialysis and Covid 19 patients admitted to hospitalization by discharge condition.

| Variable | Alive |  | Deceased |  | *p* |
| --- | --- | --- | --- | --- | --- |
|  | n | % | n | % |  |
| **N°** | 84 | 80 | 21 | 20 |  |
| **Age** | 57.65±14.47 |  | 63.5±13.24 |  | 0.19 |
| **Sex** |  |  |  |  |  |
| Female | 39 | 82.98 | 8 | 17.02 | 0.65 |
| Male | 45 | 77.59 | 13 | 22.41 |  |
| **Modality of admission** |  |  |  |  |  |
| New | 31 | 77.5 | 9 | 22.5 | 0.64 |
| Follower | 53 | 81.54 | 12 | 18.46 |  |
| **Smoker** |  |  |  |  |  |
| No | 43 | 76.79 | 13 | 23.21 | 0.354 |
| Yes | 3 | 60 | 2 | 40 | 0.258 |
| Former smoker | 38 | 86.37 | 6 | 13.63 | 0.154 |
| **CKD etiology** |  |  |  |  |  |
| HTA | 44 | 78.58 | 12 | 21.42 | 0.66 |
| no HTA | 40 | 81.64 | 9 | 18.36 |  |
| **BMI** | 24.57±2.9 |  | 24.29±3.65 |  | 0.7 |
| **Hospital stay (days)** | 13±7.63 |  | 9±6.74 |  | 0.01 |

**Table 3:** Laboratory characteristic of the CKD5 dialysis and Covid 19 patients admitted to hospitalization by discharge condition.

| Variable | Alive | Deceased | p |
| --- | --- | --- | --- |
| **Creatinine mg/dL** | 7.22±1.07 | 7.38±1.37 | 0.57 |
| **Urea mg/dL** | 159.63±28.25 | 174±38.73 | 0.06 |
| **Leukocytes** cell count x uL | 11058.71±3798.79 | 19731.43±3767.19 | <0.001 |
| **Lymphocytes** cell count x uL | 1075.32±651.38 | 582±504.53 | <0.001 |
| **Platelets** cell count x uL | 296123.81±150833.54 | 323476.19±152799.42 | 0.26 |
| **INR** | 1.34±0.77 | 1.15±0.19 | 0.63 |
| **D-dimer mg/L** | 2.64±4.65 | 5.96±12.13 | 0,01 |
| **Ferritina mg/dL** | 976.57±470.87 | 1489.33±545.65 | <0.001 |
| **CRP mg/dL** | 9.41±5.06 | 23.58±10.13 | <0.001 |
| **LDH U/L** | 377.43±59.37 | 538.19±71.17 | <0.001 |
| **pH** | 7.33±0.08 | 7.25±0.06 | <0.001 |
| **HCO<sub>3</sub> mmol/L** | 20.88±3.34 | 17.3±2.77 | <0.001 |
| **Potassium mmol/L** | 5.0±1.07 | 4.79±1.11 | 0.45 |
| **ALT U/L** | 65.61±58.74 | 53.67±24.55 | 0.93 |
| **AST U/L** | 68.92±58.37 | 59.86±27.71 | 0.85 |
| **PaO<sub>2</sub>/FiO<sub>2</sub> mm/Hg** | 195.62±34.06 | 105.67±25.56 | <0.001 |

**LDH: Lactate deshydrogenase; CRP: C-protein reactive ALT: alanine aminotransferase; AST: aspartate aminotrasnferase; pO<sub>2</sub>/FiO<sub>2</sub> Arterial Oxygen Pressure / Inspired Oxygen Fraction**

An exploratory analysis was performed by recategorizing the variables related to inflammation and the oxygen status of the patients evaluated, establishing new critical cut-off points. Increased leukocytes >11000 [HR: 3.8 (1.2-15.12); p=0.012], lymphocytes<1000 [HR: 5.21 (1.21-22.45); p=0.027], D-dimer> 1. 5 [HR: 2.87 (1.11-7.42); p=0.029], ferritin>1000 [HR: 3.05 (1.22-7.06); p=0.001], CRP>10 [HR: 27.73 (3.71-30.83), p=0.001], LDHL>400 [HR: 23.32 (3.12-174.07); p=0.002], bicarbonate ≤ 20 [HR: 7.58 (2.53-22.69); p=0.001], and PaO_2_/FiO_2_< 150 [HR: 38.01(8.83-163.49); p=0.001], were associated with increased mortality **(Table 4)**.

**Table 4:** Laboratory characteristic of the CKD5 dialysis and Covid 19 patients, re-categorized with new critical cut-off point.

| Variable | Alive |  | Deceased |  | HR (CI 95%) | *p* |
| --- | --- | --- | --- | --- | --- | --- |
|  | n | % | n | % |  |  |
| Lymphocyte |  |  |  |  |  |  |
| >1000 | 32 | 94.12 | 2 | 5.88 | 5.21 (1.21-22.45) | 0.027 |
| ≤1000 | 52 | 73.24 | 19 | 26.76 |  |  |
| Leukocytes |  |  |  |  |  |  |
| ≤11000 | 48 | 96.0 | 2 | 4.0 | 3.8 (1.2-15.12) | 0.012 |
| >11000 | 34 | 61.82 | 21 | 38.18 |  |  |
| D-dimer |  |  |  |  |  |  |
| ≤1.5 | 49 | 89.09 | 6 | 10.91 | 2.87(1.11-7.42) | 0.029 |
| >1.5 | 35 | 70 | 15 | 30 |  |  |
| Ferritin |  |  |  |  |  |  |
| ≤1000 | 57 | 89.06 | 7 | 10.94 | 3.05(1.22-7.06) | 0.016 |
| >1000 | 27 | 65.85 | 14 | 34.15 |  |  |
| CRP |  |  |  |  |  |  |
| ≤10 | 58 | 98.31 | 1 | 1.69 | 27.73(3.71-30.83) | 0.001 |
| >10 | 26 | 56.52 | 20 | 43.48 |  |  |
| LDH |  |  |  |  |  |  |
| ≤400 | 52 | 98.11 | 1 | 1.89 | 23.32(3.12-174.07) | 0.002 |
| >400 | 32 | 61.54 | 20 | 38.46 |  |  |
| Bicarbonate |  |  |  |  |  |  |
| >20 | 56 | 93.33 | 4 | 6.67 | 7.58(2.53-22.69) | 0.001 |
| ≤20 | 28 | 62.22 | 17 | 37.78 |  |  |
| PaO<sub>2</sub>/FiO<sub>2</sub> |  |  |  |  |  |  |
| >150 | 79 | 97.53 | 2 | 2.47 | 38.01(8.83-163.49) | 0.0001 |
| ≤150 | 5 | 20.83 | 19 | 79.17 |  |  |

**LDH: Lactate deshydrogenase; CRP: C-protein reactive; pO<sub>2</sub>/FiO<sub>2</sub> Arterial Oxygen Pressure / Inspired Oxygen Fraction**

Overall survival of patients at 10 and 20 days was 86.9% and 65.3%.

In multivariate analysis only CRP >10 [HR: 10.72 (95%CI 1.4-81.58), p=0.02] and PaO_2_/FiO_2_≤ 150 [HR: 44.40 (95%CI 5.86-336.06), p=0.000] were associated with lower survival **(Table 5, Figure 1 and 2)**.

**Table 05:** Factors related to survival in CKD5 dialysis and Covid 19 patients.

| Factors | HR | ES | Z | p | IC 95% |
| --- | --- | --- | --- | --- | --- |
| **CRP**>10 mg/dl | 10.72 | 11.10 | 2.29 | 0.02 | 1.4-81.58 |
| **PaO<sub>2</sub>/FiO<sub>2</sub>** ≤150 mmHg | 44.70 | 45.85 | 3.67 | 0.000 | 5.86-336.06 |

\*Cox regression

**Figure 1:**
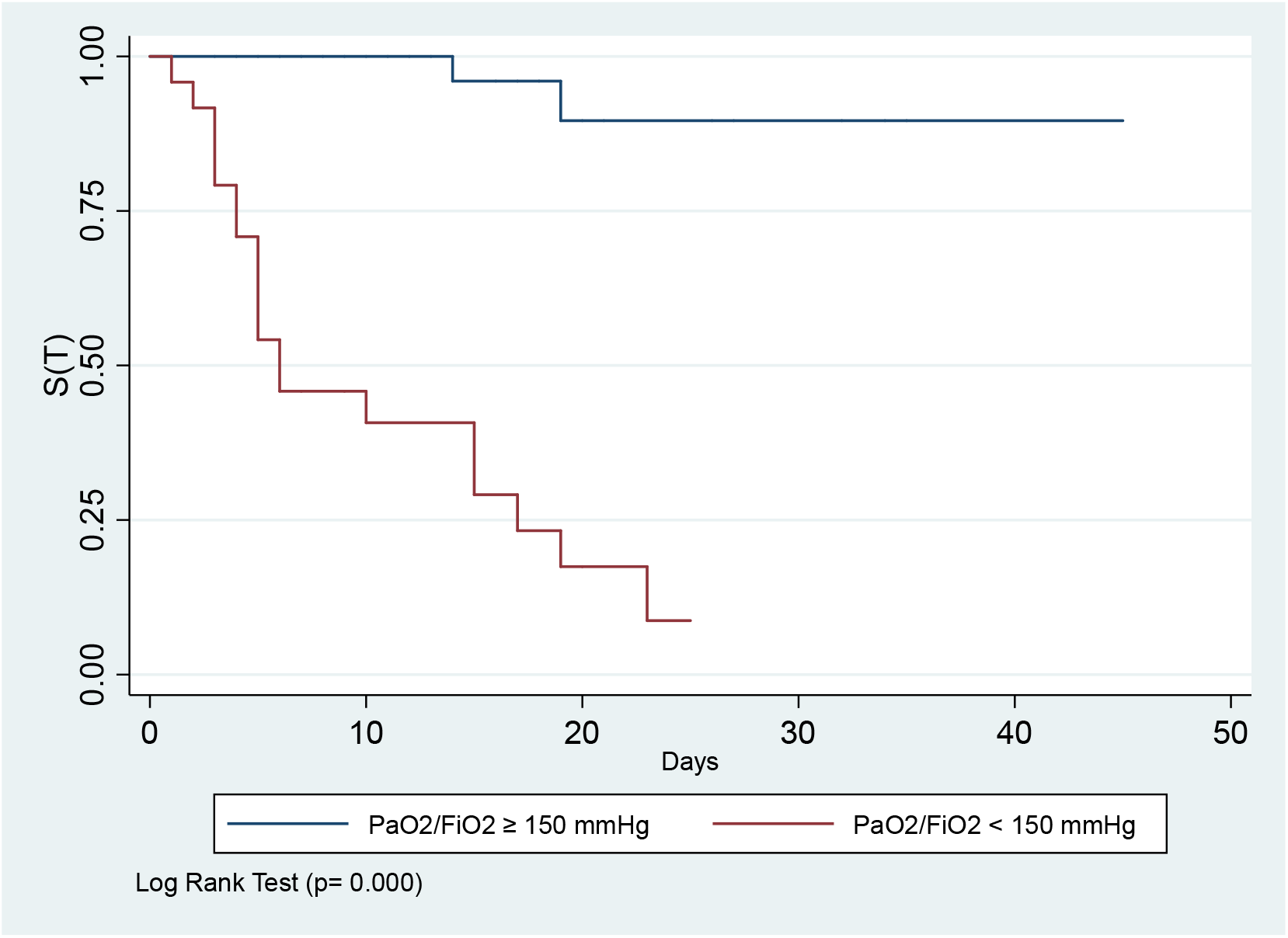
Survival in CKD5 dialysis and Covid 19 patients by level of PaO_2_/FiO_2_.

**Figure 2:**
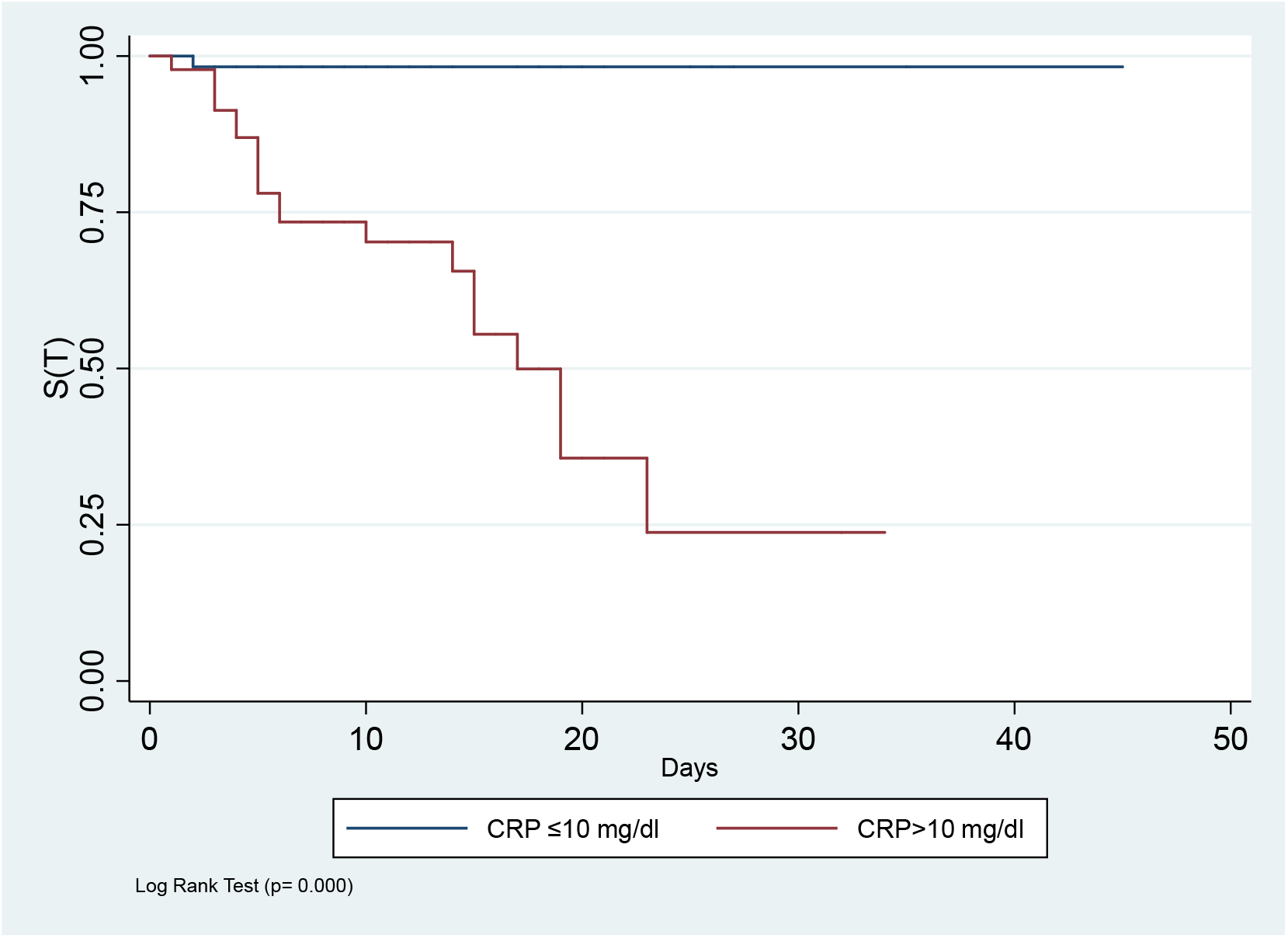
Survival in CKD5 dialysis and Covid 19 patients by level of CRP.

## DISCUSSION

It is known that the prevalence of CKD has increased in recent years in relation to the aging of the population and due to the increased prevalence of diabetes mellitus and hypertension. According to 2015 data from the Ministry of Health of Perú, kidney disease (chronic and acute) ranks seventh among the main specific causes of general mortality, with 3.3% of the total, with diabetes mellitus and arterial hypertension being the etiologies responsible for approximately 70% of the cases of Kidney Disease at the national level. Among the causes of mortality in RRT patients, the most significant were those of infectious and cardiovascular origin **^22^**. In the context of the current Covid-19 pandemic, the population of patients with stage 5 CKD in RRT also became a vulnerable population, presenting mild, moderate, and severe infection, with the need for hospitalization, requiring admission to the Intensive Care Unit (ICU) with an increase in mortality **^4,22^**.

In the present study most of the patients were male 54.29% and among the comorbidities: arterial hypertension and diabetes mellitus were the most representative with 97.14% and 23.8% of the cases respectively, similar characteristics to the general population and the population of patients with CKD infected by the Covid-19 virus **^9,10,13^**. The main cause of admission to hospitalization was respiratory failure 76.2%, requiring oxygen support in more than 80.95% of patients in the modality of reservoir mask and binasal cannula, data similar to other series **^15,22-25^**. Furthermore, Cai et al **^25^** report that between 6 - 15.5% of patients with CKD are admitted to the ICU, which differs from the present study since no patient was admitted to the ICU **^15, 22-25^**. In relation to BMI, there is a direct association between obesity and greater risk of Covid 19 infection. In the present series, the percentage of overweight was 30.48% and obesity 7.1%, lower than that reported and with no relation to mortality, this is explained because in our population on chronic dialysis obesity is less prevalent than in the general population ^15, 18, 26-28^.

The main causes of CKD were arterial hypertension and diabetes mellitus, as described in national and international reports, and the indication for dialysis support was pulmonary congestion, hyperkalemia and metabolic acidosis **^15,22-27^**, in this context, given that the symptomatic patient needed hospitalization, the time lapse between case identification, diagnosis and hospitalization was a few days, during which time the patient could receive irregular hemodialysis in outsourced centers until access to in-center dialysis, leading to manifestations of sub-dialysis and the need for emergency dialysis.

Mortality in CKD secondary to pulmonary infections in general is 51.9% and this varies according to increasing age **^18,24,27-30^**, there is evidence of reports such as that of Yang et al **^19^** on mortality in CKD infected by COVID19 which ranges between 6% and 22.7%; also in Perú a study reports up to 46% mortality **^31^**; in our research a mortality of 20% was obtained, which correlates with the aforementioned.

The baseline Charlson index, which relates long-term mortality to the patient’s comorbidity, was evaluated; at the beginning of hospitalization, it was 3.85 ±1.18 with a 10-year survival of 53.62%, taking into account that the survival rates found 20 and 30 days after hospitalization for Covid 19 infection were 65.3% and 56% respectively, it can be seen that the probability of survival decreases with hospitalization, as also reported by Savino et al **^28,30-32^**.

There is evidence that the factors related to greater mortality are age, male sex, comorbidities such as arterial hypertension, diabetes mellitus and obesity, inflammatory markers, and respiratory compromise **^25-32^**, in the present investigation as well as in a Spanish study, no relationship was found with age, sex and comorbidities **^33^**.

The severity of Covid-19 infection has not been evaluated, but it is implicitly considered that a patient hospitalized with Covid-19 is generally considered a severe infection, therefore, our study sample is made up of patients with stage 5 CKD with multiple comorbidities in addition to severe Covid-19 infection, conditions that make them a vulnerable population with high mortality.

Regarding laboratory markers, there are reports of the relationship between increased leukocytes, CRP, LDH, ferritin, D-dimer and decreased lymphocytes with mortality, data that relate the exaggerated inflammatory response in patients with Covid-19 with increased mortality, such findings are congruent with our findings **^30-33^**, in relation to the gasometrical characteristics it is described that SatO_2_<90% persistent despite increased oxygen support, pO_2_ < 68 mmHg and decreased pO_2_/FiO_2_ are related to higher mortality in the context of the severity of respiratory compromise **^34^**, in this series it was found that most patients needed oxygen support and showed a significantly decreased PaO_2_/FiO_2_. In our study the increase in inflammatory markers and indicators of oxygen status, over their values conventionally established as normal were related to mortality only in the bivariate analysis; but when categorizing these variables by establishing higher cut-off points they showed a strong relationship with a lower survival, but the variables with the greatest impact that were related to a reduction in survival were CRP values >10 mg/dl [HR: 10.72 (1.4 - 81.58) p = 0.02] and PaO_2_/FiO_2_values <150 mmHg [HR: 44.70 (5.86 - 336.06) p = 0.000], the latter also described by Wang et al **^34^**.The final results reflect that the important component related to Covid-19 mortality in hospitalized patients with CKD on chronic dialysis is the inflammatory and oxygen component.

The main limitations of the study lie in the type of design; it is a retrospective case series whose primary source of data is the hospital medical records, which implicitly may entail a series of errors in the validity and reliability of the data recorded. The biochemical and blood gas variables were processed by the hospital laboratory; they could also have a great intrinsic variability of the values since the variability of the observers and of the equipment used to process the blood samples is unknown.

## CONCLUSION

CRP levels >10 mg/dl and PaO_2_/FiO_2_≤ 150 mmHg are the main factors related to mortality and lower survival in patients with stage 5 CKD hospitalized for Covid-19.

## Data Availability

All data are available in the investigator's database in the Nephrology Unit.

## DISCLOSURE

All the authors declared no competing interests.

## FUNDING

Self-financing

## Notes

### Competing Interest Statement

The authors have declared no competing interest.

### Funding Statement

This study did not receive any type of financial support.

### Author Declarations

The Institutional Ethics and Research Committee of the Cayetano Heredia Peruvian University approved the study under the number 039-01-21 and since it is a retrospective study, it is exempt from informed consent. NOTE: At the end of the manuscript, the document of The Institutional Ethics and Research Committee

